# Clinical profiles and risk factors of hepatocellular carcinoma with viral hepatitis at a tertiary hospital in Mogadishu

**DOI:** 10.1101/2022.11.12.22282247

**Authors:** Mohammed A Hassan, Ahmed M Bashir, Isaiah G Akuku, Janet Evelia, Ahmed Aweis

## Abstract

**Objectives:** To describe the clinical profiles of HCC patients with or without viral hepatitis. Additionally, to evaluate the risk factors for HCC with viral hepatitis.

**Methods:** In a cross-sectional study, 52 HCC patients diagnosed by imaging in Mogadishu were enrolled. Serum liver blood samples were taken, and sociodemographic and risk data recorded. In addition, we used logistic regression to evaluate relevant sociodemographic and clinical risk factors for HCC with viral hepatitis.

**Results:** Of all patients, 86.5% were men, a median age of 64.0 years (interquartile range [IQR]: 51.5 to 72.0). 50% of the patients were cases of HBV or HCV, 44.2% was HBV and 15.4% were HCV. Diabetes, fatty liver disease and underweight was present in 23.1%, 30.8%, and 32.7%, respectively. The median of Gamma-glutamyl transferase (GGT) was 176.0 U/L (IQR 106.0–302.2 U/L; normal 1-60 U/L) and alpha-fetoprotein was 1337.5 ng/ml (IQR 566–2625; normal 0-8.78 ng/ml). Aspartate aminotransferase (AST) (121.5; IQR 64.5–198.0; normal range 0-31U/L), alkaline phosphatase (ALP) (173.5; IQR 125.2–232.5; normal 30-155 U/L) and direct bilirubin 0.5 (0.2–0.9; normal 0.01-0.4mg/dl) were also elevated. AST (median greatest dimension 164.5 U/L versus 70.0 U/L; p<0.001) and ALT (55.0 versus 30.0; p=0.048), and the distribution of international normalized ratio (INR) was higher (69.2% versus 34.6%; p=0.025) in viral hepatitis-positive patients than the non-viral group. Adjusted analysis showed INR was associated with viral hepatitis-HCC.

**Conclusion:** Most HCC patients were old and presented with HBV infection. Age, AST, ALT and INR biomarkers appeared to be influencing factors of viral hepatitis with HCC. The coexistence of viral hepatitis and metabolic factors enhanced the HCC development. In the primary care setting, evaluating the clinical profile could support risk stratification for HCC patients, including viral hepatitis-positive patients, timely detection, and decision-making in patient management.

## Introduction

Liver cancer is one of the leading causes of cancer-related deaths globally, accounting for 905,677 new cases and 830,180 deaths in 2020 (1). Hepatocellular carcinoma (HCC), also referred to as primary liver cancer, is the most common type of liver cancer and Africa’s fourth most common cancer (2). Viral hepatitis is a major public health issue that affects hundreds of millions of people worldwide (3). There are currently five recognized viral hepatitis types that include hepatitis A, B, C, D, E, and G viruses, with Hepatitis B Virus (HBV) and Hepatitis C Virus (HCV) strongly associated with HCC accounting for 80% of its development (3–8).

In Africa, HBV and HCV are the primary HCC aetiologic agents (2,9,10), with over half of HCC cases caused by HBV, while approximately 25% are caused by HCV (10); however, in some African countries, Egypt in particular, HCV is the leading cause of HCC (11). Unfortunately, in Sub-Saharan Africa, most HCC diagnosis is made late in the disease cycle when surgery cannot be done. Therefore, primary HCC prevention is reliant on preventing and treating viral hepatitis, especially HBV and HCV, and other underlying aetiologies (12).

Apart from the viral agents, the other important risk factors for HCC are metabolic factors (diabetes, obesity-related non-alcoholic steatohepatitis [NASH], non-alcoholic fatty liver disease [NAFLD], hereditary haemochromatosis). Additionally, carcinogenic or toxic factors (cigarette smoking, alcohol, and aflatoxins exposures) and immune-related factors (primary biliary cirrhosis or cholangitis, and 1-antitrypsin deficiency and autoimmune hepatitis) (13,14). These have been well studied outside sub-Saharan African.

Unfortunately, when HCC symptoms are manifested, the disease is in an advanced stage with poor prognosis, despite that the disease has a prolonged sub-clinical duration which provides a greater opportunity to detect the disease early for potential cure. (15). Ultrasonography, computed tomography (CT) and magnetic resonance imaging (MRI) are the commonly used imaging to identify the size of liver nodules in HCC, and tumor markers such as Alpha-fetoprotein since invasive method of biopsy might cause bleeding(16). It is also shown that alpha-fetoprotein is more sensitive in detecting HCC early in pre-clinical stage than the other tumor biomarkers such as Des-gamma carboxy-prothrombin (DCP) and lectin-bound AFP (AFP-L3)(15).

HCC is more frequent in Somalia (17), with a pooled prevalence of 14% to 29% (18); however, there is a scarcity of reliable or up to date data on the clinical and risk profile data for HBV- or HCV-related HCC in the country. Moreover, the HCC surveillance rates are also disappointingly nonexistent or low in Somalia, affecting clinical screening uptake (19). Therefore, exploring the clinical profiles and risk factors in Somalia may be useful in providing a theoretical and a practical foundation for early detection, curative options, preventing, and controlling HCC in the country. In addition, identifying markers for early diagnosis and risk stratification might benefit HCC patients in Somalia and inform a patient-centered research agenda. Therefore, in a cross-sectional study of HCC patients in Mogadishu, Somalia, we described the clinical profiles of HCC patients with or without viral hepatitis. Additionally, we explored the risk factors for HCC with viral hepatitis.

## Methods

### Study Setting and population

This was a cross-sectional study of 52 conveniently sampled patients diagnosed with HCC through imaging and laboratory test between August 2020 to August 2021 at Mogadishu Somali-Turkey Recep Tayyip Erdoğan Training and Research Hospital, Mogadishu, Somalia. This hospital, which opened its doors in Mogadishu, Banadir region, in 2015, is the country’s best-equipped referral hospital in terms of medical technology. The hospital offers tertiary care and is equipped for surgical operations and imaging procedures, such as computed tomography, ultrasound, magnetic resonance imaging, and image-guided biopsies. We excluded patients who did not provide informed consent to participate and those suffering from other severely debilitating illnesses, ostensibly frail for unknown reasons and thus unable to communicate and patients who did not provide informed consent to participate.

### Study procedures and blood sampling

All enrolled participants were interviewed using a structured questionnaire and sociodemographic data such as (age in years, sex, marital status, education, occupation, and average monthly income [United States Dollars]). We also gathered anthropometric measures (weight in kilograms and height in meters). In addition, we collected clinical history data (time since HCC was first diagnosed, HCC diagnosis method, history of HCC hospitalization, smoking status, smoking duration, family history of HCC, history of diabetes, history of fatty liver, history of physician-diagnosed liver disease, and markers of serum liver activity (Aspartate aminotransferase [AST], alanine aminotransferase [ALT], Alkaline phosphatase[ALP], Gamma-glutamyl transferase [GGT], International normalized ratio [INR] and Alpha-fetoprotein [AFP]) were collected. The tests were performed on a Mindray analyzer (Mindray) and captured.

### Blood sampling

We obtained blood samples from participants through standard venipuncture methods. In addition, we performed biochemistry tests, which included assessing serum activity levels/profiles, HBV, and HCV.

### Study definitions

Viral hepatitis was defined as a mono-infection with either HBV or HCV or an HBV/HCV co-infection. The dual infection may occur and even persist in the same patient since they have similar transmission routes (20). Body mass index (BMI) was calculated from measured height and weight and classified as normal weight (18.5–24.9 kg/m^2^), overweight (25–29.9 kg/m^2^), and obese (≥ 30 kg/m2). Current or past smoking history was self-reported by participants. Diabetes mellitus was defined as self-reported type 1 or 2 diabetes mellitus. Serum liver chemistries including AST (normal range, 0-31U/L), ALT (normal range, 0-45U/L), ALP (normal range, 30-155 U/L), GGT (normal range, 1-60 U/L), Total bilirubin (normal range, 0.30-1.10mg/dl), direct bilirubin (normal range, 0.01-0.4mg/dl), INR (normal range, 0.8-1.2.) and AFP (normal range, 0-8.78ng/ml) values were reported, and abnormal/elevated values were defined as any parameter with a value greater than the upper limit of the normal range. The AST, ALT, ALP, GGT, TBIL, DBIL, INR, and AFP threshold values were chosen based on the normal ranges used at MSTH.

### Ethics statement

The Mogadishu Somali-Turkey Recep Tayyip Erdoğan Training and Research Hospital (MSTH) ethics committee reviewed and approved the study in accordance with the Helsinki Declaration. All participants provided written informed consent before participation (28.072020, Reference number – MSTH/4125) and data were analyzed anonymously.

### Statistical Analysis

Sociodemographic, clinical variables and markers of serum liver activity levels were described overall and by HBV or HCV status since virtually all patients had elevated alpha-fetoprotein (AFP) and gamma-glutamyl transferase (GGT) above indicated cut-off level. The Shapiro Wilk test was used to test the normality of the continuous data. The continuous variables were then presented as median and their respective interquartile range (IQR: Q1 to Q3), while the categorical variables were described as frequencies and percentages. Statistical differences between groups were assessed using Fisher’s exact test for categorical variables and Mann-Whitney U two-sample test or Kruskal-Wallis test for continuous data, appropriately.

In further analysis, we evaluated the risk factors of HCC with viral hepatitis using a logistic regression model considering the sociodemographic, clinical, and aflatoxin-related factors. We used *p*-value=0.2 as a threshold for both covariate entry and exiting the final model and added clinically important covariates based on past literature. Results were expressed as adjusted odds ratio (AOR) and their 95% confidence intervals (CI). All the analyses were carried out in R (Version 4.1.2) and reported at a 5% level of statistical significance.

## Results

### Profiles of study participants

Our study included 52 patients whom 75% (n=39 was diagnosed with dual imaging of Ultrasound and CT Scan, while 13.5% (n=7) and 11.5% (n=6) diagnosed with Ultrasound and CT scan, respectively. The participants median age was 64.0 (interquartile range [IQR]: 51.5 to 72.0). Males were 45 (86.5%) while females were seven. Neither of the patients in our study was HIV-positive nor consumed alcohol. Infection with viral hepatitis was in 26 patients (50.0%), while separately HBV and HCV were in 44.2% (n=23) and 15.4% (n=8), respectively, by serology or physician diagnosis. HBV/HCV uninfected patients were older than HBV/HCV infected patients (median greatest dimension, 70.0 versus 55.0; p=0.046). Table 1 summarises the sociodemographic characteristics of all patients and stratified by HBV or HCV infection.

**Table 1.** Sociodemographic profiles of hepatocellular carcinoma patients stratified by hepatitis B virus and/or Hepatitis C virus infection

| Characteristics<sup>a</sup> | Levels | All HCC patients | HCC with HBV and/or HCV | HCC without HBV and/or HCV | *p*-value<sup>b</sup> |
| --- | --- | --- | --- | --- | --- |
| Total N (%) |  | 52 | 26 (50.0) | 26 (50.0) |  |
| Age (years) | Median (IQR<sup>c</sup>) | 64.0 (51.5 to 72.0) | 55.0 (49.8 to 69.5) | 70.0 (57.5 to 77.5) | **0.046** |
| Gender, % | Male | 45 (86.5) | 21 (80.8) | 24 (92.3) | 0.419 |
|  | Female | 7 (13.5) | 5 (19.2) | 2 (7.7) |  |
| Marital status, % | Single | 4 (7.7) | 2 (7.7) | 2 (7.7) | 0.609 |
|  | Married | 29 (55.8) | 13 (50.0) | 16 (61.5) |  |
|  | Divorced | 5 (9.6) | 4 (15.4) | 1 (3.8) |  |
|  | Widowed | 14 (26.9) | 7 (26.9) | 7 (26.9) |  |
| Education, % | Illiterate | 4 (7.7) | 3 (11.5) | 1 (3.8) | 0.671 |
|  | Quran | 29 (55.8) | 12 (46.2) | 17 (65.4) |  |
|  | Elementary | 11 (21.2) | 6 (23.1) | 5 (19.2) |  |
|  | Secondary | 2 (3.8) | 1 (3.8) | 1 (3.8) |  |
|  | Degree | 6 (11.5) | 4 (15.4) | 2 (7.7) |  |
| Occupation, % | Working | 17 (32.7) | 10 (38.5) | 7 (26.9) | 0.555 |
|  | Nonworking | 35 (67.3) | 16 (61.5) | 19 (73.1) |  |
| Average Monthly Income (USD), % | <100\$ | 25 (48.1) | 12 (46.2) | 13 (50.0) | 1.000 |
| | 100-500\$ | 22 (42.3) | 11 (42.3) | 11 (42.3) | |
| | >500\$ | 5 (9.6) | 3 (11.5) | 2 (7.7) | |
| Weight (Kg) | Median (IQR) | 55.5 (50.0 to 60.0) | 54.5 (50.0 to 60.0) | 56.0 (52.0 to 60.0) | 0.912 |
| Height (M) | Median (IQR) | 1.7 (1.6 to 1.7) | 1.7 (1.6 to 1.7) | 1.7 (1.6 to 1.7) | 0.664 |
| BMI (Kg/M<sup>2</sup>) | Median (IQR) | 16.9 (15.0 to 18.0) | 16.9 (14.7 to 18.0) | 16.4 (15.1 to 17.9) | 0.680 |
| BMI<sup>d</sup>, % | <18.5 | 17 (32.7) | 11 (42.3) | 6 (23.1) | 0.247 |
|  | 18.5–24.9 | 30 (57.7) | 12 (46.2) | 18 (69.2) |  |
|  | 25–29 | 5 (9.6) | 3 (11.5) | 2 (7.7) |  |

<sup>a</sup> N represents the total sample size for the group. USD (\$), United States Dollars; Kg, Kilograms; M, Metres; BMI, Body Mass Index. HCC, hepatocellular carcinoma; HBV, hepatitis B virus; HCV, hepatitis C virus.
<sup>b</sup> p-value from Chi-square test or Fisher's exact test for categorical variables and unpaired two-sample Wilcoxon test for continuous variables, two-sided; bold p-values indicate statistical significance (p<0.05).
<sup>c</sup> IQR, Interquartile range.
<sup>d</sup> Body Mass Index (BMI) <18.5 (Underweight), BMI 18.5–24.9 (Normal), BMI 25–29 (Overweight).

### Clinical and laboratory characteristics of HCC patients with and without HBV or HCV

Table 2 lists the clinical and laboratory findings of the patients. Since HCC was diagnosed mainly, the time was less than one year (96.2%) and was similar in both categories of HCC patients. Most of the patients reported they had no or did not know of a family history of HCC (90.4%), while 98.1% were never smokers. Twelve (23.1%) patients had a history of diabetes, and 16 (30.8%) had a history of Fatty Liver or physician-diagnosed liver disease.

**Table 2.** Clinical and laboratory characteristics of HCC patients with and without HBV and/or HCV

| Variable<sup>a</sup> | Level | All HCC patients<br>[No. of patients<br>(%) or Median<br>(Q1 to Q3)] | HCC with HBV<br>and/or HCV<br>[No. of patients<br>(%) or Median (Q1<br>to Q3)] | HCC without HBV<br>and/or HCV<br>[No. of patients (%) or<br>Median (Q1 to Q3)] | *p*-value<sup>b</sup> |
| --- | --- | --- | --- | --- | --- |
| Total N (%) |  | 52 | 26 (50.0) | 26 (50.0) |  |
| Time since<br>HCC first<br>diagnosed, % | <1 year | 50 (96.2) | 25 (96.2) | 25 (96.2) | 1.000 |
|  | 1-5 year | 1 (1.9) | 1 (3.8) | 0 (0.0) |  |
|  | >5 year | 1 (1.9) | 0 (0.0) | 1 (3.8) |  |
| History of<br>HCC<br>hospitalization<br>, % | Yes | 1 (1.9) | 1 (3.8) | 0 (0.0) | 1.000 |
|  | No | 51 (98.1) | 25 (96.2) | 26 (100.0) |  |
| Smoking<br>status, % | Yes | 1 (1.9) | 0 (0.0) | 1 (3.8) | 1.000 |
|  | No | 51 (98.1) | 26 (100.0) | 25 (96.2) |  |
| Smoking<br>duration, % | 5-10 years | 1 (1.9) | 1 (3.8) | 0 (0.0) | 0.333 |
|  | >10 years | 2 (3.8) | 0 (0.0) | 2 (7.7) |  |
| Family history<br>of HCC, % | Yes | 5 (9.6) | 1 (3.8) | 4 (15.4) | 0.262 |
|  | No | 25 (48.1) | 15 (57.7) | 10 (38.5) |  |
|  | Don't<br>know | 22 (42.3) | 10 (38.5) | 12 (46.2) |  |
| History of Diabetes, % | Yes | 12 (23.1) | 5 (19.2) | 7 (26.9) | 0.743 |
|  | No | 40 (76.9) | 21 (80.8) | 19 (73.1) |  |
| Fatty Liver Disease<sup>c</sup> | Yes | 16 (30.8) | 11 (42.3) | 5 (19.2) | 0.132 |
|  | No | 36 (69.2) | 15 (57.7) | 21 (80.8) |  |
| AST | Median (IQR) | 121.5 (64.5 to 198.0) | 164.5 (123.8 to 288.2) | 70.0 (45.2 to 116.0) | **<0.001** |
|  | Normal (0-31U/L) | 6 (11.5) | 1 (3.8) | 5 (19.2) | 0.191 |
|  | Abnormal | 46 (88.5) | 25 (96.2) | 21 (80.8) |  |
| ALT | Median (IQR) | 39.0 (23.0 to 65.8) | 55.0 (35.0 to 80.8) | 30.0 (20.0 to 58.8) | **0.048** |
|  | Normal (0-45U/L) | 28 (53.8) | 10 (38.5) | 18 (69.2) | 0.050 |
|  | Abnormal | 24 (46.2) | 16 (61.5) | 8 (30.8) |  |
| ALP | Median (IQR) | 173.5 (125.2 to 232.5) | 191.0 (147.0 to 218.0) | 157.0 (98.2 to 284.2) | 0.481 |
|  | Normal (30-155 U/L) | 21 (40.4) | 8 (30.8) | 13 (50.0) | 0.258 |
|  | Abnormal | 31 (59.6) | 18 (69.2) | 13 (50.0) |  |
| GGT<sup>2</sup> | Median (IQR) | 176.0 (106.0 to 302.2) | 206.0 (132.0 to 327.2) | 132.5 (82.0 to 200.8) | 0.151 |
|  | Normal (1-60 U/L) | 5 (9.6) | 2 (7.7) | 3 (11.5) | 1.000 |
|  | Abnormal | 47 (90.4) | 24 (92.3) | 23 (88.5) |  |
| Total bilirubin | Median (IQR) | 0.7 (0.5 to 1.4) | 1.1 (0.5 to 1.6) | 0.6 (0.4 to 1.1) | 0.050 |
|  | Normal (0.30-1.10mg/dl) | 34 (65.4) | 14 (53.8) | 20 (76.9) | 0.144 |
|  | Abnormal | 18 (34.6) | 12 (46.2) | 6 (23.1) |  |
| Direct bilirubin | Median (IQR) | 0.5 (0.2 to 0.9) | 0.5 (0.4 to 0.9) | 0.3 (0.2 to 0.8) | 0.156 |
|  | Normal (0.01-0.4mg/dl) | 24 (46.2) | 9 (34.6) | 15 (57.7) | 0.164 |
|  | Abnormal | 28 (53.8) | 17 (65.4) | 11 (42.3) |  |
| INR | Median (IQR) | 1.2 (1.0 to 1.6) | 1.4 (1.2 to 2.0) | 1.1 (1.0 to 1.3) | **0.001** |
|  | Normal (0.8-1.2) | 25 (48.1) | 8 (30.8) | 17 (65.4) | **0.025** |
|  | Abnormal | 27 (51.9) | 18 (69.2) | 9 (34.6) |  |
| AFP | Median (IQR) | 1337.5 (566 to 2625) | 884.0 (460.2 to 3600) | 1614 (1052 to 2261) | 0.558 |
|  | Normal (0-8.78ng/ml) | 2 (3.8) | 1 (3.8) | 1 (3.8) | 1.000 |
|  | Abnormal | 50 (96.2) | 25 (96.2) | 25 (96.2) |  |

<sup>a</sup> Body Mass Index (BMI) <18.5 (Underweight), BMI 18.5–24.9 (Normal), BMI 25–29 (Overweight); HCC, hepatocellular carcinoma; HBV, hepatitis B virus; HCV, hepatitis C virus; AST, Aspartate aminotransferase; ALT, Alanine aminotransferase; ALP, Alkaline phosphatase; GGT, Gamma-glutamyl transferase; INR, International normalised ratio; AFP, Alpha-fetoprotein.
<sup>b</sup> p-value from Chi-square test or Fisher's exact test for categorical variables and unpaired two-sample Wilcoxon/Mann-Whitney U test for continuous variables, two-sided; bold p-values indicate statistical significance (p<0.05).
<sup>c</sup> Fatty Liver Disease – ever been diagnosed with fatty liver or ever been told by a doctor you have liver disease.

Nearly all the serum levels were elevated for the markers. The overall median of GGT and AFP was 176.0 (106.0 to 302.2) and 1337.5 (566 to 2625), respectively but were statistically indifferent when stratified by HBV/HCV status. The median AST (greatest dimension 164.5 U/L versus 70.0 U/L; p<0.001) and ALT (55.0 versus 30.0; p=0.048) were higher in patients with HBV/HCV than those without HBV/HCV. INR distribution was higher (69.2% versus 34.6%; p=0.025) in viral hepatitis-positive HCC patients than the viral hepatitis-negative HCC patients. Table 2 details the serum levels and status of HBV/HCV infection.

### Pairwise correlations for liver-related seromarkers, age and BMI

We carried out correlation analysis with continuous variables, as shown in Table 3. We observed a significant but negative correlation between age, AST and INR. Seromarker AST significantly and positively correlated with ALT, ALP, GGT, TBIL, DBIL and INR. ALT also significantly and positively correlated with ALP and GGT. We also demonstrated a positive and significant correlation for ALP with GGT, TBIL, DBIL and INR. A significant positive correlation was also found between GGT and TBIL, DBIL and INR. TBIL correlated significantly with DBIL and INR.

**Table 3.** Spearman pairwise correlations for liver biomarkers, age and BMI (n=52)

| Variables<sup>a</sup> | Age | BMI | AST | ALT | ALP | GGT | TBIL | DBIL | INR | AFP |
| --- | --- | --- | --- | --- | --- | --- | --- | --- | --- | --- |
| Age | 1 |  |  |  |  |  |  |  |  |  |
| BMI | 0.06 | 1 |  |  |  |  |  |  |  |  |
| AST | -0.30* | 0.08 | 1 |  |  |  |  |  |  |  |
| ALT | 0.05 | 0.12 | 0.61* | 1 |  |  |  |  |  |  |
| ALP | 0.14 | -0.16 | 0.32* | 0.52* | 1 |  |  |  |  |  |
| GGT | -0.08 | -0.10 | 0.44* | 0.33* | 0.63* | 1 |  |  |  |  |
| TBIL | 0.00 | -0.14 | 0.37* | 0.24 | 0.30* | 0.35* | 1 |  |  |  |
| DBIL | 0.07 | -0.14 | 0.36* | 0.24 | 0.35* | 0.33* | 0.75* | 1 |  |  |
| INR | -0.31* | -0.21 | 0.31* | 0.13 | 0.29* | 0.49* | 0.31* | 0.21 | 1 |  |
| AFP | 0.02 | -0.22 | -0.21 | -0.13 | 0.11 | -0.05 | 0.00 | 0.20 | 0.14 | 1 |

<sup>a</sup> Spearman correlations ( $\rho$ ) were done for age, AST (aspartate aminotransferase), ALT (alanine aminotransferase), ALP (alkaline phosphatase), GGT (gamma glutamyl transferase), INR (international normalized ratio), AFP (alpha-fetoprotein), BMI (body mass index), and TBIL (total bilirubin), DBIL (direct bilirubin).
\* Indicates statistically significant correlation/relationship.

### Multivariable analysis of risk factors for hepatocellular carcinoma in patients with hepatitis B or C virus

A logistic regression model assessed the risk factors for HCC with HBV or HCV. The model’s calibrative-ability (Hosmer-Lemeshow Test Chi-sq (8) 8.37, p=0.398), and discriminative-ability (C-statistic = 0.822) was good. Findings show that a unit increase in INR in the univariable model (odds ratio [OR] 14.64, 95% confidence interval [CI] 3.07-120.9, p=0.004) and in the multivariable model (adjusted OR 10.61, 95% CI 1.80-120.13, p=0.024) was associated with HCC in patients with hepatitis B or C virus. Age, family history of HCC, BMI, and fatty liver disease were all statistically attenuated (p>0.05). The results are summarised in Table 4.

**Table 4.** Risk factors for hepatocellular carcinoma in patients with hepatitis B or C virus

| Variable<sup>a</sup> | Level | HCC with HBV and/or HCV | HCC without HBV and/or HCV | Logistic regression<sup>c</sup> |  |
| --- | --- | --- | --- | --- | --- |
|  |  |  |  | Univariable | Multivariable |
| | | | | OR (95% CI, $p$-value) | AOR (95% CI, $p$-value) |
| Age (years) | Median (IQR)<sup>b</sup> | 55.0 (49.8 to 69.5) | 70.0 (57.5 to 77.5) | 0.97 (0.93-1.00, $p=0.089$) | 0.98 (0.94-1.03, $p=0.407$) |
| BMI (kg/m<sup>2</sup>) | BMI 18.5–24.9 | 12 (40.0) | 18 (60.0) | Reference | Reference |
| | BMI <18.5 | 11 (64.7) | 6 (35.3) | 2.75 (0.82-9.95, $p=0.108$) | 1.61 (0.31-8.15, $p=0.558$) |
|  | BMI 25–29 | 3 (60.0) | 2 (40.0) | 2.25<br>(0.33-19.10,<br>p=0.411) | 2.75<br>(0.23-42.43,<br>p=0.426) |
| HCC Family history | Absent | 25 (53.2) | 22 (46.8) | Reference | Reference |
|  | Present | 1 (20.0) | 4 (80.0) | 0.22<br>(0.01-1.63,<br>p=0.190) | 0.16<br>(0.01-1.46,<br>p=0.152) |
| Fatty Liver Disease<sup>b</sup> | No | 15 (41.7) | 21 (58.3) | Reference | Reference |
|  | Yes | 11 (68.8) | 5 (31.2) | 3.08<br>(0.92-11.55,<br>p=0.077) | 1.83<br>(0.39-9.00,<br>p=0.443) |
| INR | Median (IQR) | 1.4 (1.2 to 2.0) | 1.1 (1.0 to 1.3) | 14.64<br>(3.07-120.9,<br>p=**0.004**) | 10.61<br>(1.80-120.13,<br>p=**0.024**) |

<sup>a</sup> BMI, Body Mass Index <18.5 kg/m<sup>2</sup> (underweight), BMI 18.5–24.9 kg/m<sup>2</sup> (Normal), BMI 25–29 kg/m<sup>2</sup> (Overweight); HCC, hepatocellular carcinoma; HBV, hepatitis B virus; HCV, hepatitis C virus; INR, international normalized ratio; IQR, interquartile range (Q1 to Q3).
<sup>b</sup> Fatty Liver Disease – ever been diagnosed with fatty liver or ever been told by a doctor you have liver disease.
<sup>c</sup> The model fitted using logistic regression for hepatocellular carcinoma with Hepatitis B or C. Number of observations = 52, Number observations in model = 52, Missing = 0, AIC = 67.5, C-statistic = 0.822, Hosmer-Lemeshow Test= Chi-sq (8) 8.37 (p=0.398). Variables/factors associated with p-values <0.2 in univariable analysis were entered into a multivariable logistic regression analysis. OR, odds ratio; AOR, adjusted odds ratio; CI, confidence interval. Bold p-values indicate statistical significance (p<0.05).

## Discussion

We described the clinical profiles of HCC patients with or without viral hepatitis and evaluated the risk factors for HCC with viral hepatitis. We described viral hepatitis as mono- or co-infection with HBV and HCV. We observed clinically significant elevated AST, ALP, GGT, direct bilirubin and AFP serum liver chemistries. We also found that nearly all patients were non-smokers, less than 10% had a family history of HCC, a fifth were diabetic, and less than a third had a history of fatty liver diseases. Furthermore, we established that INR was positively associated with viral hepatitis–positive HCC.

Of the two hepatic viral infections in our study, HBV was the predominantly common type (44.2%), and the mono- and co-infections explained HCC in 50% of the study population. However, a considerable proportion (30.8%) of the patients had a history of fatty liver/ a physician-diagnosed liver disease was a prime aetiology of HCC in this patient group, especially in terms of non-alcoholic fatty liver and NASH (inflammation, liver damage, and fat accumulation in the liver), which are all forms of NAFLD. Moreover, like in our study, the coexistence of NAFLD and viral hepatitis, particularly HBV, has been observed frequently(21), sometimes with an inverse association(22,23). We observed that 42.3% of viral hepatitis–positive HCC patients reported fatty liver disease. Such concurrent fatty liver has been said to increase the risk of virus-associated HCC (24–26) and potentially lead to cancer in our study population. Additionally, none of the patients in our study was HIV-positive, eliminating possible synergistic interaction between HBV or HCV and HIV.

Diabetes and HCC, like other cancers, are both multifarious, distinct, and long-term, fatal diseases that have a significant impact on health(27). Our study found that diabetes was moderately prevalent and had no statistically significant association with viral hepatitis–positive HCC. This finding affirms previous studies that reported that diabetes, as a risk factor for HCC, exists independently of the availability of HBV or HCV, non-specific cirrhosis, or alcoholic liver disease (28–31).

Our study observed that the HCC patients were considerably old (median 64.0 years; IQR, 51.5 to 72.0; range, 25 to 90) but appeared comparable to the mean of 52.6 years reported in a recent retrospective Somalian study (32). Ageing is a known risk factor for HCC development (33,34), as it impairs liver function and decreases the liver’s capacity to regenerate (35). We also observed that viral hepatitis–negative HCC patients were much older than the viral hepatitis group and were mostly diabetic. The previous description may be explained by NAFLD, which studies have previously reported to be associated with ageing, diabetes and concurrent viral hepatitis infection (24–26,36).

In our study, most of the patients had normal BMI (18.5–24.9 Kg/M^2^), and none were obese. Although obesity and diabetes are risk factors for HCC and may vary according to HBV or HCV infection status (37), as with diabetes, we did not observe a difference in BMI distribution by HBV or HCV infection status in our study. However, a considerable number of patients were underweight (32.7%).

We found that AST, ALP, GGT, direct bilirubin, INR, and AFP serum levels were markedly elevated in most of the patients in our study population. A third (34.6%) and less than half (46.2%) had elevated total bilirubin and ALT. Additionally, slightly more than half had elevated direct bilirubin (53.8%) and INR (51.9%). Elevated liver enzymes in older people are a common finding and are often linked to metabolic syndrome(33,35,38). The serum levels for GGT, AST, ALT, and ALP, appear similar to previous Saudi Arabian findings(39). It is worth mentioning that 96.2% of the patients in this study had elevated AFP levels which is a sign of poor prognosis. A study by Asaoka et al. had shown how the elevated tumor markers including AFP are important predictors of vascular invasion that can suggest metastasis of the primary tumor and is a poor prognosis.(40)

Evidence demonstrates that cigarette smoking is associated with increased progression and severity of liver disease, particularly fibrosis and liver cancer (41–43). However, in our study, none of the patients reported consuming alcohol. Furthermore, only one reported smoking cigarette, thus eliminating the potential contribution of alcoholic liver disease to or significant effect of cigarette smoking on HCC development. Additionally, we observed that the patient who reported smoking cigarettes had elevated GGT, ALP and INR but were viral hepatitis–negative. For this patient, cigarette smoking may have had a carcinogenic or an inflammatory effect on the liver (44). This is corroborated by Wannamethee and Shaper(45), who reported a significant association between cigarette smoking with increased GGT and ALP levels but not with high AST levels.

The most frequently encountered serum liver enzyme levels in clinical practice are hepatocellular predominance with increased ALT and AST and cholestatic predominance with elevated ALP and GGT (46). AST and ALT are released into the bloodstream by damaged hepatocytes, and their activities have long been recognised as sensitive indicators of liver disease (47–49). Still, they may also reflect non-specific hepatocellular damage(50). Elevated ALT levels have also been associated with severe liver damage and serve as a diagnostic biomarker for NAFLD (51). Overall, we found a positive correlation between AST and ALT. We also observed a significant difference in the distribution of AST and ALT by viral infection status, and this could be due to acute/chronic viral hepatitis or NASH, implying that AST and ALT may be used as prognostic or predictive biomarkers of viral hepatitis–positive HCC in this study population.

In our study, we observed abnormally higher values of GGT and ALP in viral hepatitis–positive HCC patients than in the viral hepatitis–negative HCC group; however, the difference was not statistically significant. This could be due to cholestasis or biliary obstruction, which would result in elevated GGT and ALP due to impaired bile formation, secretion, or flow. The observed elevated GGT enzyme serum levels in over 90% of the patients appeared to predict the development of HCC in our overall study population.

Serum AFP levels were elevated in over 95% of the patients and stratified by virus-infection status. Our finding is inconsistent with Giannini et al.(52)’s Italian study that had elevated AFP levels in 62.4% of HCC patients. AFP is a biomarker whose serum concentration rises in most patients with HCC; consequently, it has been utilised in the monitoring and diagnosis of or early detection of neoplasms in HCC (53,54), despite its contested role (55). It may not be an indispensable diagnostic or screening tool (56); however, our study showed that serum AFP levels were markedly elevated. Similarly, GGT has also been proposed as a potential marker for the early detection of HCC. Furthermore, the levels of the two biomarkers can also be used to determine the survival outcome; that is, the higher the levels of GGT and AFP, the poorer the survival outcome (57). However, we observed no clear linear relationship between AFP and GGT, implying that these two biomarkers are likely associated with distinct HCC characteristics (58) in our study population.

HCC is frequently diagnosed in patients with pre-existing liver cirrhosis, but it can also develop without such a condition. Liver cirrhosis and high serum ALT are two factors that may influence serum AFP elevation (56). Although we did not test liver cirrhosis in our study, we did not find a clear correlation between AFP and ALT. The interaction between ALT levels and HCC onset and development has influences serum AFP levels (56). However, nearly all the HCC patients with elevated AFP presented elevated GGT levels simultaneously.

We observed higher levels of coagulation marker (INR) in viral hepatitis-positive patients than in the viral hepatitis-negative group. The World Health Organization developed the INR standardised reporting of prothrombin time coagulation function markers (59). The INR has been used to standardise the PT value in liver diseases. The liver is essential in the synthesis of blood coagulation factors and, thus, the importance of checking levels of INR. A high level of INR indicates slower clotting of blood, hence a higher risk of prolonged bleeding.

This is among the first few studies in Somalia that provides profile and risk data for viral hepatitis-related HCC. The strength of our study is that the viral hepatitis-positive and viral hepatitis-negative groups had a balanced number of patients and detailed data on routinely tested liver enzymes and seromarkers. However, this study had several limitations. Firstly, our study had a single-centre cross-sectional design hence prognosis was largely unfeasible as the biomarkers were not measured dynamically. Secondly, some liver-or HCC-related data or co-factors that could influence the findings were not collected, such as the aflatoxins test and our inability to identify the medications used by the patients. Thirdly, some findings were merely patient-reported and not verifiable by medical records, indicating possible recall bias. Lastly, the sample size was relatively small compared to the burden of viral hepatitis in Somalia (16), which makes it not a good representation; hence generalizability to other settings remains to be evaluated – a major limitation of our study.

In conclusion, most HCC patients present with HBV infection and are considerably old. AST, ALP, GGT, direct bilirubin, INR, and AFP serum levels were markedly elevated in most HCC patients. Age, AST, ALT and INR biomarkers appeared to be influencing factors of viral hepatitis with HCC. The HCC development appeared to have been enhanced by the coexistence of viral hepatitis and metabolic factors. In the primary care setting, evaluating the clinical profile could support risk stratification for HCC patients, including viral hepatitis-positive patients, timely detection and decision-making in patient management.

## Data Availability

All data produced in the present study are available upon reasonable request to the authors

## Acknowledgement

We express our sincere appreciation to Musab Ahmed Nor, a Physician at Somali-Turkey Recep Tayyip Erdoğan Training and Research Hospital who has assisted us in the data collection.

